# Characterization of the suicide attempt-associated chromosome 7 locus

**DOI:** 10.1101/2025.07.13.25331392

**Authors:** Laura M. Fiori, Anjali Chawla, Vikram Nathan, Malosree Maitra, Corina Nagy, Gustavo Turecki

## Abstract

Suicide is one of the leading causes of death worldwide. Although suicidal behaviors demonstrate high heritability, identifying the underlying genetic factors has been challenging. Recent genome-wide association studies for suicidal behavior identified a SNP on chromosome 7, rs62474683, which was the most significantly associated SNP. As this SNP is intergenic, the mechanism by which it may be related to suicidal behaviors is unclear. In order to determine the potential functional effects of the rs62474683 genotype, and how it may be related to suicidal behavior, we ascertained expression of genes within a 1.8Mbp region surrounding this SNP in two brain regions in individuals who died by suicide, and investigated the relationship between genetic variation and gene expression. Additionally, we explored, at the single cell level, the effect of the variant on gene expression and chromatin accessibility. While we found several genes displaying differential expression, Forkhead box P2 (FOXP2) was the most consistently altered in brains of individuals who died by suicide. and its expression was related to rs62474683 genotype. Furthermore, the relationship between FOXP2 expression and suicide appeared to be both brain region- and cell type-specific. Finally, we found evidence for a direct relationship between the region containing rs62474683 and FOXP2, and identified Homeobox family transcription factors as potential mediators of the relationship between FOXP2 and the rs62474683 SNP. In conclusion, our study provides evidence for a functional relationship between the most significant suicide attempt-associated locus to date and genes displaying differential expression in individuals who died by suicide.

## Introduction

Suicidal behaviors range from passive suicidal ideation to death by suicide. Suicide accounts for over 727,000 deaths worldwide each year ^1^, with significantly higher numbers of individuals attempting suicide or experiencing suicidal ideation. The majority of individuals exhibiting suicidal behaviors have a history of psychiatric disorders ^2, 3^, however, risk for suicide appears to be transmitted independently of the liability for psychopathology ^4^.

Numerous twin ^5^, adoption ^6^, and family ^7, 8^ studies have demonstrated that suicidal behaviors possess a genetic component, with heritability estimated between 30-50% ^4, 9–11^, with the remainder of the risk due to environmental factors. Numerous biological pathways have been implicated in suicidal behaviors, including stress response systems, monoaminergic neurotransmission, and inflammation. However, candidate gene association studies investigating these systems have not been able to explain the high heritability of suicidal behaviors. Suicidal behaviors are complex phenotypes, and genetic vulnerability involves differential contributions from numerous genes and biological pathways. Recently, a large genome-wide association study (GWAS) of suicidal behavior, which comprised data from 29,782 individuals with histories of suicide attempts and/or death by suicide, and 519,961 controls, was performed by the International Suicide Genetics Consortium (ISGC) ^12^. This study identified two loci which were related to suicidal behavior at the genome-wide level: the major histocompatibility complex (MHC), and an intergenic locus on chromosome 7. The association between the chromosome 7 SNP, rs62474683, and suicidal behavior, was replicated in the independent Million Veterans Program (MVP) cohort. Interestingly, the association did not appear to be mediated by risk for psychiatric disorders, indicating it is specifically associated with suicidal behavior. A follow-up study involving a multi-ancestry GWAS meta-analysis of the ISGC and MVP studies (comprising 43,871 individuals with suicide attempts or death by suicide, and 915,025 ancestry-matched controls) identified eight genome-wide significant loci, with rs62474683 displaying the strongest association with suicidal behavior ^13^. The functional implications of this SNP are not known, as it lies within a large intergenic region, with the closest gene being a long non-coding RNA, *LINC01392*, 149kb upstream.

Our goal in the present study was to functionally characterize relationships between rs62474683 and nearby genes, and to determine their relevance to suicidal behavior. We identified several nearby genes which were differentially expressed in the brains of individuals who died by suicide, and found that expression of two of these genes was influenced by rs62474683 genotype. Moreover, using chromatin profiling, we found evidence for direct relationships between the linkage disequilibrium (LD) block containing rs62474683, and LD blocks containing expression quantitative trait loci (eQTLs) for several of these genes. Our study thus provides evidence for a functional relationship between the most significant suicidal behavior risk locus to date and genes displaying differential expression in the brains of individuals who died by suicide.

## Materials and methods

### Human samples

Post-mortem samples of dorsal ACC (Brodmann area (BA) 24) and dorsolateral prefrontal cortex (DLPFC, BA8/9) were obtained from the Douglas-Bell Canada Brain Bank (Douglas Mental Health University Institute, Montreal, Quebec, Canada). Sample characteristics are shown in Supplementary Table 1. Psychological autopsies were performed as described previously, based on DSM-5 criteria ^14^. The control group had no history of major psychiatric disorders. All cases died by suicide and met criteria for major depressive disorder (MDD) or depressive disorder not-otherwise-specified. The majority of subjects were of French-Canadian origin, a population with a known founder effect ^15^. Written informed consent was obtained from next-of-kin. This study was approved by our local institutional review board.

### Gene expression

#### RT-PCR

RNA was extracted from bulk tissue using the miRNeasy Mini kit (Qiagen). Quality of RNA was assessed using the Agilent 2200 Tapestation, and only samples with RNA Integrity Number (RIN) ≥ 5.0 were used. RNA was reverse-transcribed using M-MLV Reverse Transcriptase (ThermoFisher) with a combination of oligo (dT) and random primers. RT-PCR was performed using SYBR green (Applied Biosystems) with GAPDH as an endogenous control using primers shown in Supplementary Table 2. Similar results were found using the geometric mean of GAPDH and β-actin (not shown). Reactions were run in triplicate using the QuantStudio 6 Flex System, and data was collected using QuantStudio Real-Time PCR Software v1.1. Expression levels were calculated using the absolute (standard curve method) quantification method. Differential expression was assessed using t-tests or Mann-Whitney tests, after removal of outliers. Differential gene expression findings remained significant after controlling for age and RIN.

#### Single-nucleus RNA-sequencing (snRNA-seq)

This sample was previously described in ^16^. Briefly, we ran high-throughput snRNA-seq on the 10X Genomics Chromium platform in 38 female subjects from the DLPFC. Libraries were prepared with the Chromium Single Cell 3’ Reagent Kits version 3.1 or 2. Additionally, we simultaneously re-processed a previously published DLPFC snRNA-seq dataset (GSE144136) in males (N=33) ^17^. Subjects are described in Table 1. Alignments and gene counts were generated with Cell ranger version 5.1.0 (10X Genomics). We used the highly variable genes across nuclei to perform PCA, corrected for batch effects with the Harmony R package ^18^, and performed unsupervised clustering of nuclei using the Seurat R package ^19^. Clusters were annotated to specific cell types based on enrichment of expression of known marker genes. The average expression of all genes in all nuclei from each subject within each broad cell type was calculated, scaling each nucleus to 10000 unique molecular identifiers (UMIs). Gene expression was transformed, and group comparisons between broad cell clusters were performed using Mann-Whitney tests. Additionally, a sub-cluster analysis for FOXP2 was performed using a combination p-value approach, as described in ^16^.

### Genotyping

#### rs62474683

Genotyping of rs62474683 was performed using a Taqman assay (C 88877036_10). DNA was diluted to 20ng/µL, and analysed in duplicate using the using the QuantStudio 6 Flex System and data was collected using QuantStudio Real-Time PCR Software v1.1.

#### Genotyping array

We genotyped a sample of primarily French-Canadian individuals using the Illumina Global Screening array including the Psych 30K panel, which was performed at Genome Quebec. Data was exported from Genome Studio and processed in PLINK. Quality was assessed by analysing call rate, unexpected relatedness, sex mismatches, and Hardy-Weinberg equilibrium (hwe). After removing low quality samples, we had genotyping information for 134 individuals (68 C, 66 S). Variants with minor allele frequency (MAF) < 1%, call rate < 95%, or hwe p-value < 0.000001 were removed. Imputation was done with the following procedure: SHAPEIT was used to phase variants for each subject, using the full 1000 Genomes phase 3 dataset, then IMPUTE2 was used to impute new variants using the haplotypes produced by SHAPEIT. The 1000 Genomes phase 3 dataset was used as the reference panel. Imputation was performed across 3 Mbp intervals on each chromosome, with effective population size (Ne) of 20,000 and 500 reference haplotypes selected for each individual. After imputation, variants that had info score < 0.8, MAF < 0.01, or hwe p-value < 0.000001 were removed. 8,821,074 variants remained after these filters.

#### LD blocks and eQTL analysis

Using the imputed dataset for 134 subjects, we identified LD blocks for the entire chromosome 7 using PLINK, following the definitions found in ^20^. Coordinates are based on hg19. We performed an eQTL analysis for DLPFC expression (RT-PCR) using variants within 500kb of FOXP2, MDFIC, TFEC, and TES, after post-imputation filtering. matrixEQTL ^21^ was used for the analysis, and two PCs generated with PLINK’s --pca flag (EIGENSTRAT with default settings) were includes as covariates. The "modelANOVA" option was used to test for additive and dominant effects.

### Chromatin interactions

#### Single Nucleus Assay for Transposase-Accessible Chromatin-seq (snATAC-seq)

The collection and processing of snATAC-seq data has been previously described in (https://www.biorxiv.org/content/10.1101/2023.10.02.560567v1.abstract). Briefly, we performed snATAC-seq in 84 samples from the DLPFC (40C, 44S), described in Table 1. Nuclei were extracted from homogenized brain samples, purified, and captured using the 10X platform. Sequencing was performed at Genome Quebec using the Illumina NovaSeq 6000. Processing and quality analyses of sequencing data was performed using ArchR (release_1.0.1) ^22^.

#### Peak-to-Peak Connections

We used the DLPFC snATAC-seq data from approximately 200k cells to identify peak-to-peak co-accessibility between peaks (501bp) and peak-to-gene expression correlations (using sample-matched snRNA-seq data) within 1 Mbp windows using the ArchR pipeline ^22^.

#### Histone modifications

Neuronal (NeuN+) and non-neuronal (NeuN-) DLPFC histone modification data ^23^ was collected from Synapse and converted to hg38 coordinates using UCSC liftOver tool (v377).

#### Hi-C Connections

Neuronal and non-neuronal HiC DLPFC data ^24^ was also converted to hg38 coordinates. Hi-C connections were assessed by overlapping them with chromosome 7 regions of interest.

#### S-LDSC

We computed the enrichment of GWAS SNPs associated with suicide attempt ^12^ in the open chromatin regions (OCRs) of each DLPFC cell-type using a stratified LDSC regression model (v 1.0.1). DLPFC cell-type OCRs (501bp) were identified using MACS2 (q<0.1) ^25^ and used for S-LDSC analysis as previously described [https://www.biorxiv.org/content/10.1101/2023.10.02.560567v1.abstract]. Briefly, OCRs were converted from hg38 to hg19 coordinates using the UCSC liftover tool (v377). Cell-type specific partitioned heritability analysis was computed using a full baseline-LD model and adding LD scores estimated on respective OCR sets combined across all cell types as background. One-sided p-values were computed based on the enrichment z-scores for each annotation relative to the background.

#### Transcription factor (TF) analyses

We used the motifbreakR package ^26^ (method=”ic”, filterp = TRUE, threshold = 1e-4) with the SwissRegulon position weight matrix (PWM) from the MotifDb package to evaluate TF motifs potentially disrupted by the rs62474683 variant. The resulting p-value was calculated using calculatePvalue (granularity = 1e-6) and variant effect with “strong” disruption was selected. Additionally, Homer ^27^ was used to identify TF motifs enriched in the OCRs associated with FOXP2 LD block 4, with default parameters.

## Results

### Expression of genes near rs62474683 in the ACC and DLPFC of individuals who died by suicide

In order to identify functional effects of rs62474683 genotypes that may be related to suicidal behavior, we focused on genes surrounding this SNP, including Forkhead box P2 (FOXP2), MyoD family inhibitor domain containing (MDFIC), LINC01393, LINC01392, transcription factor EC (TFEC), and Testin LIM domain protein (TES), spanning a 1.8 Mbp region.

We first assessed the expression of these genes in bulk postmortem ACC and DLPFC tissue obtained from individuals who died by suicide and compared them to neurotypical controls (Figures 1a and 1b). In the ACC, FOXP2 displayed significantly upregulated expression in individuals who died by suicide (p=0.019), whereas TFEC and TES were significantly downregulated (p=0.044 and 0.020, respectively). In the DLPFC, both FOXP2 and TES were downregulated in individuals who died by suicide (p=0.025 and 0.005, respectively). There were no differences in MDFIC expression in either brain region. The expression of LINC01393 was too low to be accurately quantified in the majority of our samples (not shown), while LINC01392 was undetectable.

**Figure 1:**
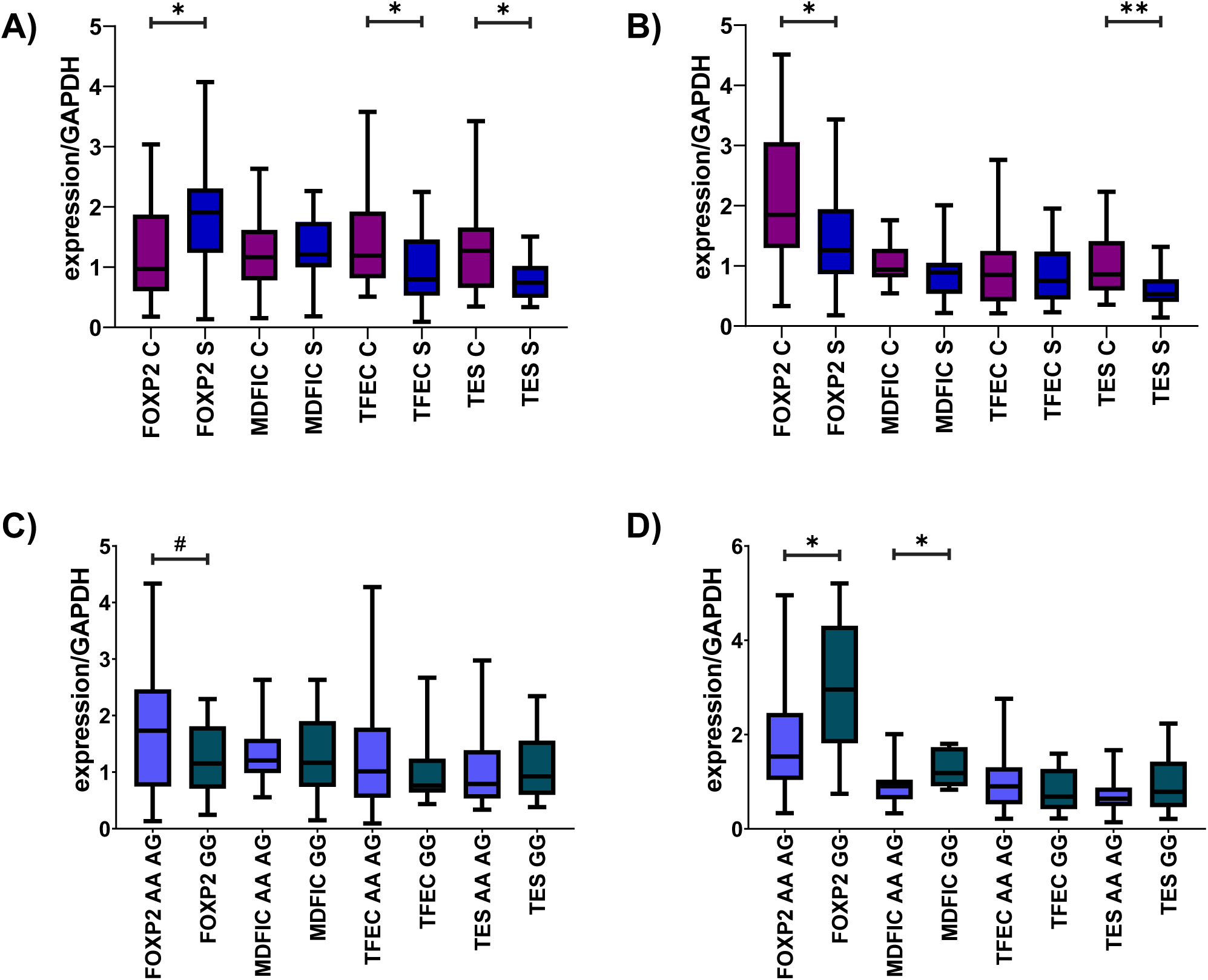
Gene expression in the ACC (A) and the DLPFC (B) and relationship between rs62474683 and gene expression in the ACC (C) and the DLPFC (D). RT-PCR was performed in neurotypical controls (C) (purple), and individuals who died by suicide (S) (blue). Expression was compared between carriers of the A allele (AA and AG, blue) and GG genotype (turquoise).

### Relationship between expression in the brain and rs62474683

We next wanted to determine if rs62474683 variation was associated with brain expression of the genes in the candidate region. To this end, we genotyped rs62474683. As the A allele represented the risk allele for suicidal behavior ^12^, for each gene, we combined individuals with AA and AG genotypes and compared them to those with the GG genotype. In both the ACC and DLPFC, genotype influenced the expression of FOXP2, such that individuals with the A allele had a trend for higher expression in the ACC (Figure 1c, p=0.055), whereas in the DLPFC, A allele carriers displayed lower expression (Figure 1d, p=0.031). Additionally, carriers for the A allele had lower expression of MDFIC in the DLPFC (p=0.010). We further compared the expression levels of FOXP2 and MDFIC between the three rs62474683 genotypes (Supplementary Figure 1). In the DLPFC, there were significant differences between FOXP2 AA and GG genotypes (p=0.05), MDFIC AG and GG genotypes (p=0.04), and a trend for a significant difference between MDFIC AA and GG genotypes (p=0.07). The ACC showed no overall significant differences between genotypes (p=0.13 and 0.84 for FOXP2 and MDFIC, respectively).

### Cell-type specific expression in the DLPFC

Given that FOXP2 displayed both gene expression differences related to suicide, and demonstrated an association between gene expression and rs62474683 variation, we focused the remainder of our analyses on examining the expression patterns of this gene, and how FOXP2 expression may be related to rs62474683 genotypes. To better understand the different cellular contributions to the associations we observed between expression and suicidal behavior, we assessed the cell type specific expression of FOXP2 in the DLPFC, using single-cell transcriptomic data we previously generated in individuals who were depressed and died by suicide and are publicly available ^16^ across seven broad cell types (astrocytes, endothelial cells, excitatory neurons, inhibitory neurons, microglia, oligodendrocytes, and oligodendrocyte precursor cells). Results are shown in Figure 2 and Supplementary Figure 2. We found several clusters displaying differential expression of FOXP2, with the most significantly decreased expression in excitatory neurons (p=0.003) in individuals who died by suicide and increased expression in astrocytes (p=0.036), oligodendrocytes (p=0.006), and with a trend in oligodendrocyte precursor cells (p=0.081). We further assessed the expression of FOXP2 at the sub-cell cluster level (Figure 2b), and again the most significant differences were found in excitatory neurons (subcluster ExN4_L35), followed by inhibitory neuron subcluster InN5_SST, and oligodendrocyte precursor cell subcluster 2. Expression levels of MDFIC, TFEC, and TES are shown in Supplementary Figure 2, and demonstrate that TES displayed increased expression in excitatory neurons (p=0.034), while no differences were observed for MDFIC or TFEC, which is consistent with overall expression differences in bulk DLPFC tissue.

**Figure 2:**
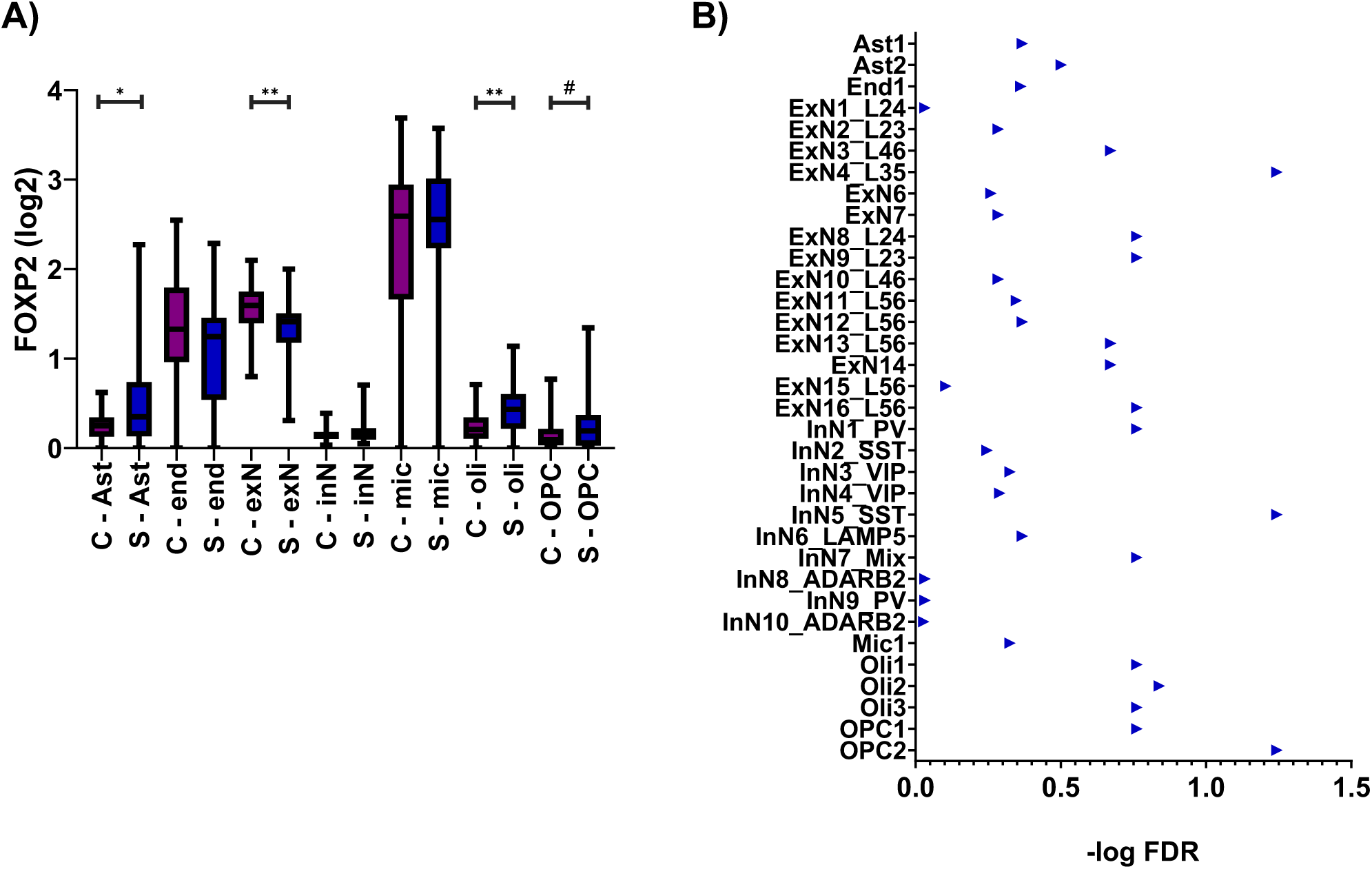
Differential expression of FOXP2 in relation to suicide in the DLPFC. A) Single-cell RNA-sequencing results are shown for astrocytes (Ast), endothelial cells (end), excitatory neurons (exN), inhibitory neurons (inN), microglia (mic), oligodendrocytes (oli), and oligodendrocyte precursor cells (OPC). Mann-Whitney tests were used for group comparisons between controls (C, purple) and individuals who died by suicide (S, blue). B) Results from combination p-value analyses for differential expression of FOXP2 at the sub-cell cluster level.

### eQTL analysis in the DLPFC and relationship with rs62474683

In order to determine how the intergenic rs62474683 may be related to expression of FOXP2, we first performed an eQTL analysis for this gene in the DLPFC. Results are shown in Figure 3 and Supplementary Tables 3-5. We identified 12 SNPs within 500kb of FOXP2 which were nominally associated with its expression. We then identified the LD blocks containing these cis eQTLs using the LD blocks generated in a larger sample of individuals from the same population, and identified 5 LD blocks containing cis eQTLs, and 4 eQTL SNPs which were not annotated to LD blocks. Two of these LD blocks (FOXP2 LD blocks 3 and 4), and one SNP (rs5886733) were shared eQTLs with MDFIC. Similar analyses were performed for MDFIC, TFEC, and TES, and are shown in Supplementary Tables 3-5 and Supplementary Figures 3-5. In total, we identified 44 LD blocks containing eQTLs (FOXP2: 5, MDFIC: 22, TFEC: 7 and TES: 10), and 67 eQTLs which were not assigned to an LD block.

**Figure 3:**
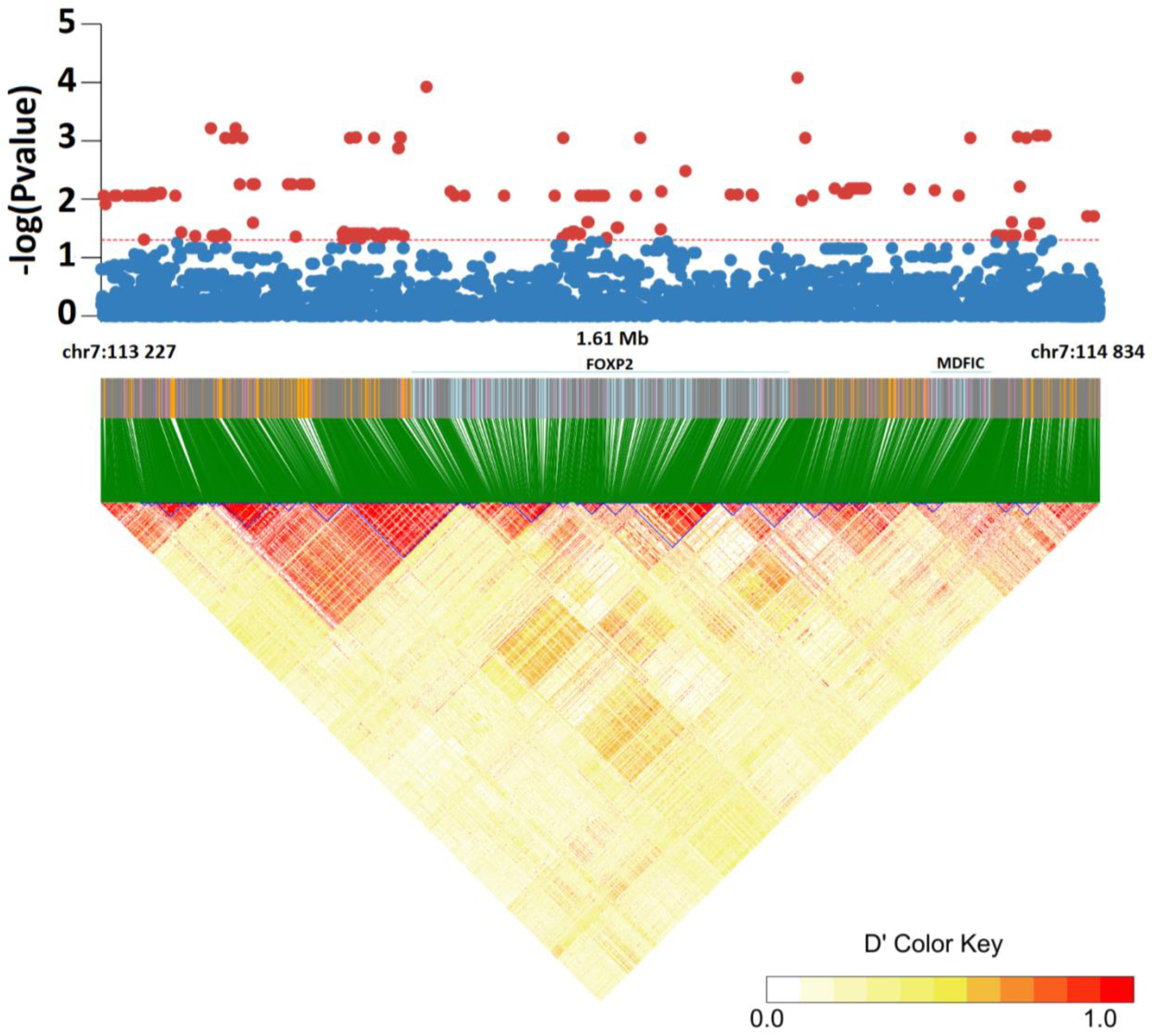
Expression quantitative trait loci (eQTL) and linkage disequilibrium (LD) block analyses for FOXP2. Upper panels indicate relationship between individual SNPs and gene expression. Lower panels indicate LD structure of the regions in a sample composed primarily of French-Canadian individuals.

### Gene regulatory relationship between rs62474683 LD region and DLPFC eQTLs

Next, in order to identify potentially functional relationships between the LD block containing the intergenic rs62474683 and LD blocks containing eQTLs, we used DLPFC snATAC-seq data we recently produced from individuals who were depressed and died by suicide (https://www.biorxiv.org/content/10.1101/2023.10.02.560567v1.abstract). Using snATAC-seq data, 60% of all FOXP2 (3/5 LD blocks) and 59% of MDFIC (13/22) eQTL LD blocks had significant open chromatin peak-to-peak connections (FDR<0.05; an indication that these regions may either be co-regulated or that one region is functionally regulating the other) with the rs62474683 LD block (Supplementary Figure 6a). Among connections with FOXP2, LD block 4 had the most significant correlation (r=0.22, FDR<0.05, peak region=chr7:114861277-114861777) with the rs62474683 LD region (Figure 4). The same peak region showed peak-to-gene correlation (FDR<0.05, an indication that open chromatin accessibility is correlated with gene expression using snRNA-seq data) with both FOXP2 (r=0.17, FDR<0.05) and MDFIC (r=0.15, FDR<0.05) and overlapped (within 500bp) with a SNP identified as an eQTL (rs10272695) for both FOXP2 and MDFIC genes (Supplementary Tables 6, 8 and 11). As this connection corresponded to one of the shared FOXP2/MDFIC LD blocks, these results suggest that the same region, found between these two genes, may regulate the expression of both genes. This is particularly interesting given that both MDFIC and FOXP2 showed the strongest relationships between expression in the DLPFC and rs62474683 genotype (Figure 1).

**Figure 4:**
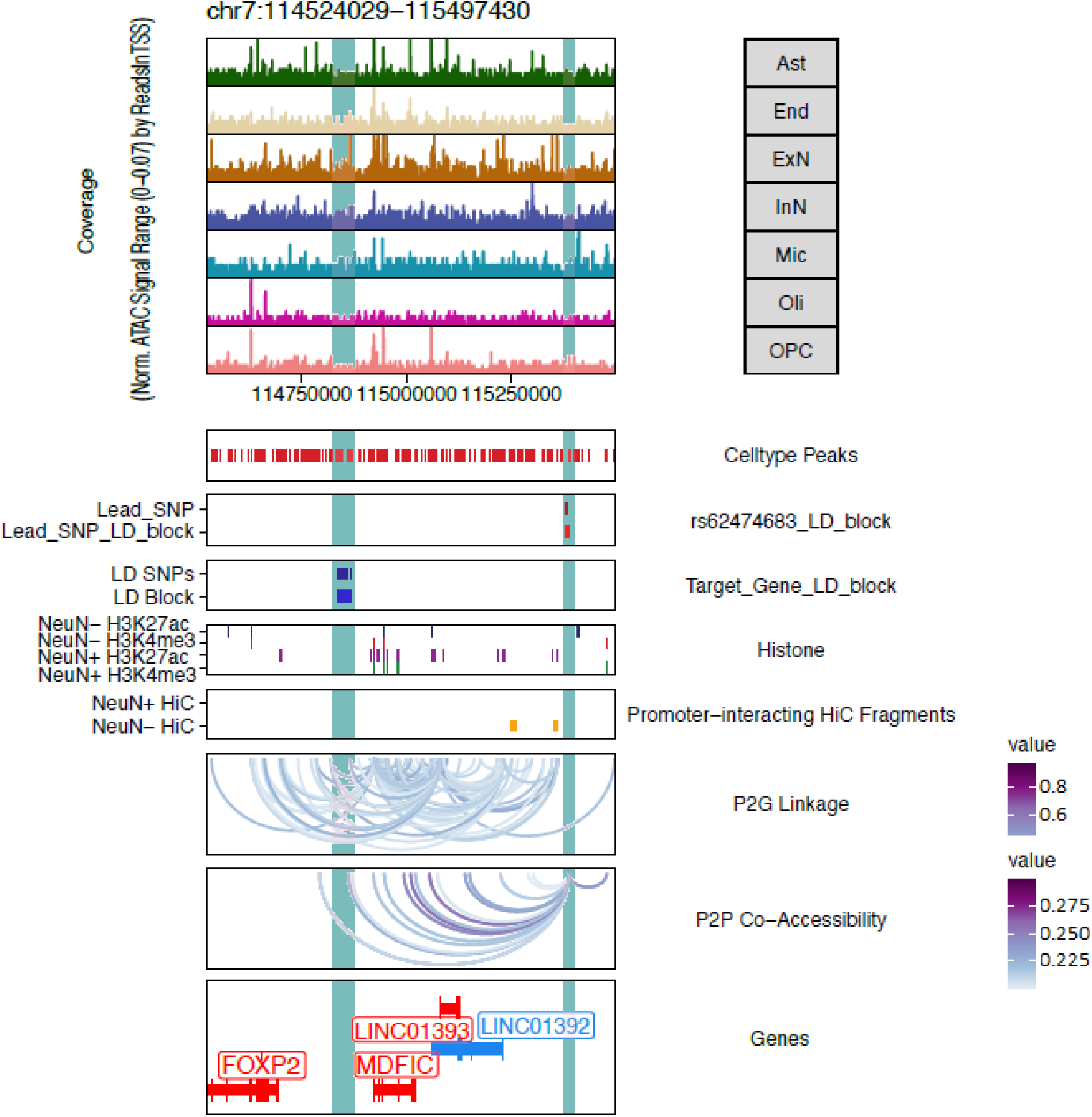
Direct peak-to-peak connection between rs62474683 and FOXP2 LD block 4. snATAC-seq based pseudobulk chromatin accessibility in DLPFC broad cell-types (Ast, End, ExN, InN, Mic, Oli, OPC); snATAC-seq open chromatin peaks; rs62474683 LD block; FOXP2 eQTL LD block 4; DLPFC neuronal and non-neuronal (NeuN+/-) histone modification peaks ^23^; DLPFC promoter-interaction NeuN+/- Hi-C fragments ^24^; Correlation of snATAC-seq Celltype Peaks with snRNA-seq expression of genes present in the “Genes” panel (Peak2GeneLinks: P2G); Co-accessibility (Peak2Peak: P2P) of FOXP2 LD block 4 with rs62474683 LD block (correlation=r=0.22, FDR<0.05).

A previous *in vitro* study ^28^ identified and characterized two shared enhancer regions for FOXP2 and MDFIC (chr7:114456873-114463136 and chr7: 114541370-114543683, hg19), which were adjacent to the shared LD 4/5 region found in the present study. Deletion of these enhancers downregulated expression of FOXP2 and upregulated expression of MDFIC, and their effects appeared to be tissue-specific, which may explain our own findings of brain-region specific relationships between rs62474683 genotype and FOXP2 and MDFIC expression. Using our paired snATAC-seq and snRNA-seq data, we confirmed that one of the previously identified enhancer regions (hg19:chr7:114816818-114823081/hg38:chr7:114456873-114463136) overlapped with four unique open chromatin regions showing significant peak-to-gene linkages (FDR<0.05) with FOXP2 and MDFIC (Supplementary Table 9), which further supports the validity of our functional gene regulatory approach.

All direct peak-to-peak connections identified between rs62474683 and eQTL LD blocks are summarized in Supplementary Tables 6 and 8. Additionally, one SNP for FOXP2 and seven for MDFIC, which were not part of LD blocks, also displayed peak-to-peak correlations within 500bp of open chromatin peaks in the rs62474683 LD block. Results are summarized in Supplementary Table 7.

We next performed a stratified linkage disequilibrium score regression (S-LDSC) using suicide attempt associated genetic variation data, and found the most significant enrichment in the open chromatin regions of excitatory neurons (Figure 5, p=0.002). As we also found a significant association between rs62474683 variation and FOXP2 gene expression in excitatory neurons (Figure 2), we confirmed the presence of a functional relationship between rs62474683 and FOXP2 LD block 4 by restricting peak-to-peak correlations within these neurons (Supplementary Figure 6b).

**Figure 5:**
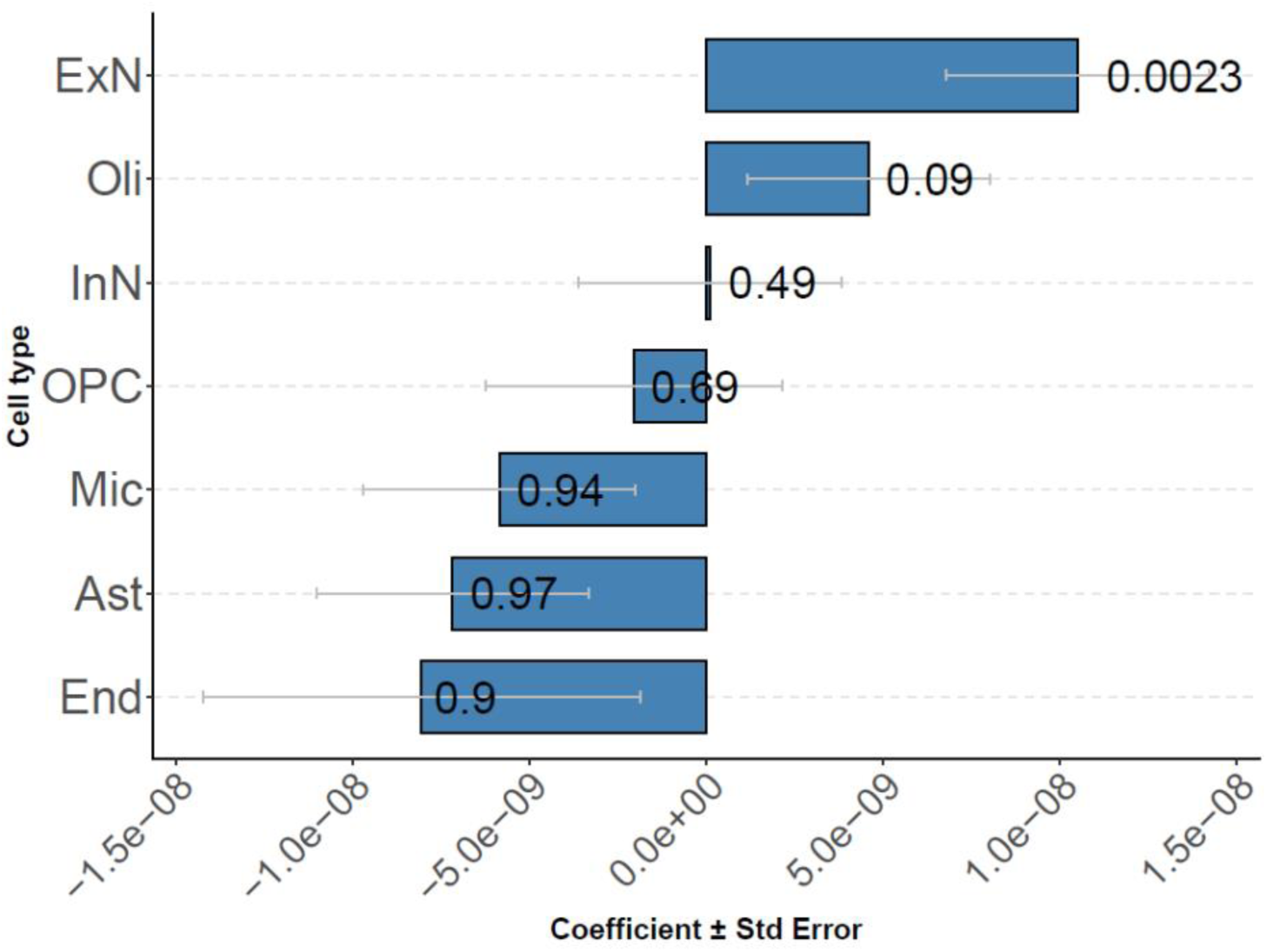
Relationship between cell type and suicide attempt-associated genetic variants: Bar plot showing the coefficient (+/- standard error) on x-axis from linkage disequilibrium score regression (LDSC) analysis of suicide attempt associated genetic variants enriched in the open chromatin regions identified in each cell type in snATAC-seq data (LDSC z-score test, p values for each cell type are displayed on the bar plot).

Further, we investigated open chromatin regions adjacent to the rs62474683 LD block (within 20 kbp) with significant peak-to-peak connections (r>0.2, FDR<0.05) with FOXP2 LD block 4. Two open chromatin peaks (including chr7:114861277-114861777 and chr7:114835793-114836293) with direct connections (Supplementary Table 8) also showed connections to a region 10kbp upstream of the rs62474683 LD block. The strongest peak-to-peak connection (r=0.41) from FOXP2 LD block 4 (chr7:114865508-114866008) was found approximately 18kbp from the rs62474683 LD block (Supplementary Figure 7). Interestingly, this chromatin region also demonstrated the strongest peak-to-gene correlation with FOXP2 (r=0.42) and MDFIC (r=0.3) and overlaps with a FOXP2/MDFIC eQTL SNP (rs2188306) (Supplementary Tables 10 and 11). Overall, our analysis suggests that rs62474683 genetic variation may be regulating FOXP2 gene expression in excitatory neurons by interacting with the open chromatin regions associated with FOXP2 LD block 4 (i.e. containing eQTLs for FOXP2) and containing an experimentally characterized enhancer.

Finally, we aimed to identify transcription factors (TFs) that may be enriched in the open chromatin regions associated with FOXP2 LD block 4, as they may be regulating FOXP2 gene expression. Motifs for TFs associated with the Homeobox family (Bapx1/Nkx3.2 and Nkx3.1) were significantly enriched in these regions (p=1e-2, Homer, Figure 6a). Interestingly, a TF from the same family (Homeobox, Nkx.6) with similar motif sequence (ttaagtGg) was found to be potentially disrupted by the rs62474683 alternate allele (G, p=9.90811e-05) compared to the reference allele (A) (Figure 6b). These TFs are known to share sequence homology and tend to function together in various developmental processes and cell fate determination ^29, 30^. Therefore, it may be possible that these TFs act together in regulating FOXP2 expression by rs62474683.

**Figure 6:**
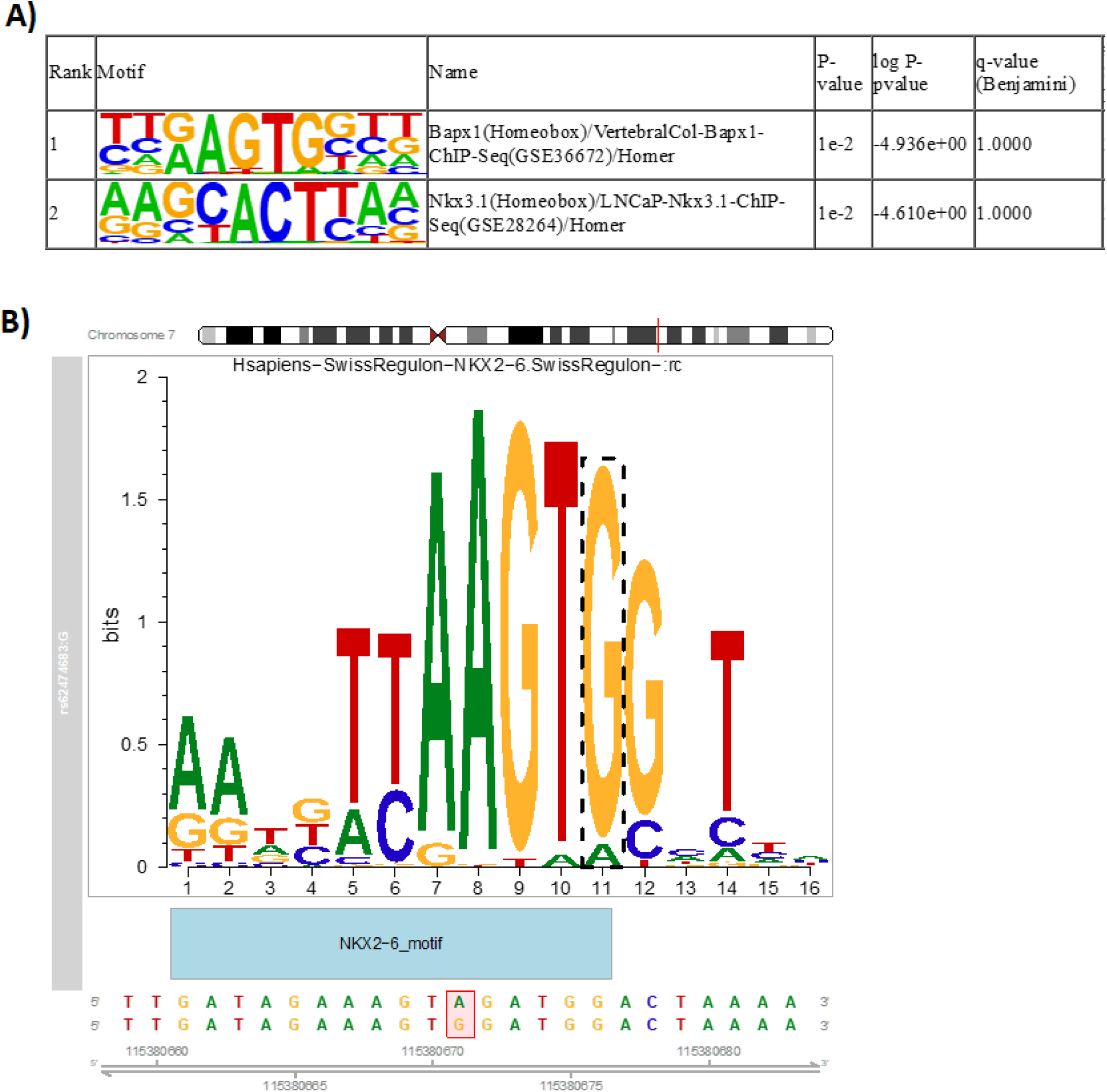
Transcription factor analysis of rs62474683 and FOXP2 regions. A) Homer was used to identify enrichment of known TF motifs (p<0.05) in the open chromatin regions associated with FOXP2 LD block 4. B) MotifbreakR plot shows the TF (NKX2-6) motif sequence logo predicted to be significantly disrupted (p=9.90811e-05) by rs62474683 alternate allele (G) highlighted in the red box below.

## Discussion

Our goal in the present study was to identify and characterize potential functional relationships between a suicide attempt-associated locus on chromosome 7, and nearby genes, and to determine how this may be related to suicide. Overall, we identified differential expression of several genes in the region surrounding rs62474683 in the brains of individuals who died by suicide, as well as a relationship between expression of some of these genes and rs62474683 genotypes. Moreover, we found evidence suggesting direct relationships between the LD block containing rs62474683, and eQTLs and LD blocks containing eQTLs for FOXP2, MDFIC, and TES.

Of the genes we investigated, FOXP2 appears to be the strongest candidate gene for mediating the relationship between rs62474683 and suicide. This gene was differentially expressed in both the ACC and DLPFC, displayed cell-specific differential expression in the DLPFC, and was influenced by rs62474683 genotype. Moreover, an eQTL-LD block for this gene appears to be functionally related to the LD block containing rs62474683, suggesting a direct, functional, relationship between rs62474683 genotype and FOXP2 expression. FOXP2 is a transcription factor which is essential for brain development, where it regulates synapse formation ^31^, neurite outgrowth ^32^, and neurogenesis ^33^. It plays a crucial role in speech and language development, and disruption of this gene has been determined to cause severe speech and language disorders (OMIM 602081) ^34^. Furthermore, gene expression differences and variants within FOXP2 have been associated with post-traumatic stress disorder ^35^, attention deficit hyperactivity disorder ^36^, schizophrenia ^37^, depression ^38, 39^, substance use disorders ^40^, cannabis use disorder ^41^, learning disorders ^42^, antisocial behaviors ^43^, and autism ^44^. Interestingly, a recent study found that FOXP2 was specifically upregulated in humans, compared to other primates, in two sub-clusters of excitatory neurons in the posterior cingulate cortex, whereas its levels were decreased in one of these sub-clusters within the prefrontal cortex and ACC ^45^. Additionally, FOXP2 has been shown to display human-specific expression in microglia, and primate-specific expression in layer 4 excitatory neurons in the DLPFC ^46^. These findings are in agreement with our own data demonstrating both cell-type specific expression of FOXP2 in the DLPFC, and contrasting results between brain regions. Furthermore, our previous analyses found FOXP2 to be a marker gene for the sub-cluster of excitatory neurons that displays the most differentially expressed genes between individuals with MDD and neurotypical controls, as well as being enriched in microglia ^17^. Differences in cell populations or regulatory mechanisms governing FOXP2 beyond those related to rs62474683 may explain the differences in directionality of suicide-related expression in the present study, both between the ACC and DLPFC, and between broad cell types within the DLPFC. A more thorough investigation of FOXP2 expression across the brain, along with expression of its gene targets is warranted.

Although we found evidence for regulation of MDFIC expression by the rs62474683 block, this gene was not differentially expressed in the ACC or DLPFC of individuals who died by suicide. MDFIC is a transcriptional regulator that modulates gene expression by sequestering transcription factors ^47^. Intriguingly, MDFIC has been shown to modulate the glucocorticoid receptor (GR) transcriptome, such that MDFIC binding to the GR is responsive to glucocorticoids, and GR downregulates MDFIC expression ^48^. This is particularly interesting given the importance of the GR and the hypothalamic-pituitary-adrenal axis in stress related disorders and suicidal behaviors, and it is possible that MDFIC expression in other tissues, or during periods of stress, may indeed be relevant for suicidal behaviors. A recent study examining genetic vulnerability to antisocial behaviors and psychiatric disorders found that SNPs mapping to MDFIC and FOXP2 influenced the shared risk for antisocial behaviors and MDD, as well as antisocial behaviors and PTSD ^49^. Additionally, differential methylation of MDFIC in cord blood was found in children exposed to prenatal depression and anxiety who were also exposed to selective serotonin reuptake inhibitors (SSRIs), compared to those with healthy mothers and those who were exposed to prenatal depression and anxiety but not SSRIs ^50^. This suggests that MDFIC expression could be influenced by antidepressant exposure, which should be assessed in future studies.

There are a number of limitations that should be noted. Firstly, all individuals who died by suicide in our sample also had a history of depression. As such, we cannot be certain that our gene expression findings relate to suicide rather than depression. Future work in cohorts of individuals with other psychiatric conditions are necessary to disentangle these effects. Secondly, our study cohort was primarily of French-Canadian origin, and it is unclear if results are generalizable to other populations. Thirdly, our study was cross-sectional, and only investigated two brain regions. Given the developmental importance of FOXP2 in particular, analysis of these genes in other brain regions, at other timepoints, will be required to better understand the breadth and timing of gene expression changes related to suicidal behaviors. Indeed, three of the genes investigated in this study, FOXP2, MDFIC, and TFEC, are transcriptional regulators which are induced under specific conditions, and may have greater importance at different periods prior to death. Furthermore, analysis of genes which are regulated by FOXP2, MDFIC, and TFEC should be performed in order to identify those which may be more directly related to suicidal behaviors. Finally, we were unable to quantify the expression of the gene which was closest to rs62474683, LINC01393, in the majority of our samples. Intriguingly, although most individuals had levels of this gene which were below the limit of detection, a few subjects displayed consistently high levels of LINC01393 across several brain regions (not shown), suggesting there may be an important genetic component to regulation of this gene. Future work in larger samples may help identify eQTLs for this gene, and their potential relationship with rs62474683.

In summary, our work has highlighted FOXP2 as the potential downstream effector of the suicide attempt associated SNP on chromosome 7, such that it displays differential expression in the brains of individuals who died by suicide, and its expression appears to be related to rs62474683 genotype through an intergenic region between FOXP2 and MDFIC. Future studies are warranted to more fully investigate the involvement of this gene in suicidal behaviors, as well as the potential for this gene to be used as a biomarker or treatment target.

## Data Availability

All data produced in the present study are available upon reasonable request to the authors

## Acknowledgements

GT holds a Canada Research Chair (Tier 1) and is supported by grants from the Canadian Institutes of Health Research (CIHR; FDN148374, PJT183903, PJT189993) and National Institutes of Health (NIH; R01MH131818). CN is a FRQS research scholar (Junior 1).

